# One side effect – two networks? Lateral and postero-medial stimulation spreads induce dysarthria in subthalamic deep brain stimulation for Parkinson’s Disease

**DOI:** 10.1101/2023.04.26.23289100

**Authors:** Hannah Jergas, Jan Niklas Petry-Schmelzer, Jonathan Hannemann, Tabea Thies, Joshua N. Strelow, Ilona Rubi-Fessen, Jana Quinting, Juan Carlos Baldermann, Doris Mücke, Gereon R. Fink, Veerle Visser-Vandewalle, Till A. Dembek, Michael T. Barbe

## Abstract

**Objective:** This study aims to shed light on structural networks associated with stimulation-induced dysarthria (SID) and to derive a data-driven model to predict SID in patients with Parkinson’s Disease (PD) and deep brain stimulation (DBS) of the subthalamic nucleus (STN).

**Methods:** Randomized, double-blinded monopolar reviews determining SID thresholds were conducted in 25 patients with PD and STN-DBS. A fiber-based mapping approach, based on the calculation of fiber-wise Odds Ratios for SID, was employed to identify the distributional pattern of SID in the STN’s vicinity. The ability of the data-driven model to classify stimulation volumes as “causing SID” or “not causing SID” was validated by calculating receiver operating characteristics (ROC) in an independent out-of-sample cohort comprising 14 patients with PD and STN-DBS.

**Results:** Local fiber-based stimulation maps showed an involvement of fibers running lateral and postero-medial to the STN in the pathogenesis of SID, independent of the investigated hemisphere. ROC-analysis in the independent out-of-sample cohort resulted in a good fit of the data-driven model for both hemispheres (AUC_left_ = 0.88, AUC_right_ = 0.88).

**Interpretation:** This study reveals an involvement of both, cerebello-thalamic fibers, as well as the pyramidal tract, in the pathogenesis of SID in STN-DBS. The results may impact future postoperative programming strategies to avoid SID in patients with PD and STN-DBS.

## Introduction

Deep brain stimulation (DBS) of the subthalamic Nucleus (STN) is a safe and effective treatment for patients with Parkinson’s Disease (PD) if medical treatment alone does not provide sufficient relief of symptom burden and motor complications.^1,2^ However, STN-DBS can induce troublesome and potentially therapy-limiting side effects, such as dysarthria. Therefore, programming strategies that avoid this disabling side effect are needed. Unlike hypokinetic dysarthria affecting up to 90% of patients with PD as per natural disease progression,^4^ DBS-related, stimulation-induced dysarthria (SID) leads to more heterogeneous speech symptom profiles with highly variable changes in intelligibility related to modulation of the speech motor system in both cross-sectional and longitudinal studies.^5–12^ The supposed pathomechanism of SID is the spread of stimulation beyond the target area, especially involving surrounding white matter tracts, such as the pyramidal tract and cerebello-thalamic fibers.^13–17^ While some studies reported a positive effect on SID by a reduction of pulse width and stimulation frequency, it seems plausible that especially proper lead positioning and shaping of the local current spread, e.g. by segmented, “directional” contacts, might be the key for avoiding SID.^18–22^

In the present prospective, double-blinded study we systematically assessed the occurrence of SID in patients with PD and bilateral STN-DBS. The clinical results were combined with a state-of-the-art fiber-based local mapping approach and validated in an independent out-of-sample cohort, to deepen our understanding of local stimulation spread associated with SID. The study aimed to support clinicians in reshaping the patient-specific current spread to avoid SID.

## Methods

### Patient Cohort and Ethics

Patients with PD and bilateral STN-DBS, implanted at least three months before study participation, were included in this prospective, double-blinded monocenter study. Additional inclusion criteria were an age between 40 and 80 years and German as the native language. Eligibility for STN-DBS was determined outside of this study by an interdisciplinary DBS-board, and based on a confirmed diagnosis of PD according to the UK brain bank criteria and the fulfillment of the CAPSIT-PD standards.^23^ The study was carried out following the Declaration of Helsinki and was approved by the local ethics committee (vote N° 20-1097). All patients gave written informed consent prior to study participation.

### Clinical Assessment

All assessments took place after an overnight withdrawal of dopaminergic medication. First, patients were assessed in “stimulation ON” with the respective clinical stimulation setting and in “stimulation OFF” after a washout period of at least 15 minutes. Before testing, impedances were checked to exclude faulty contacts from further examination. Then, a hemisphere-wise monopolar review was conducted, while stimulation of the contralateral hemisphere remained switched off. Each lead was tested at the respective 4 contact levels, with omnidirectional stimulation mode for directional levels (all Cartesia™, Boston Scientific). Default values were used for frequency (130 Hz) and pulse width (60 μs). Hemispheres and contact levels were evaluated in a randomized order to maintain blinding of the raters and the patient. During the monopolar review the amplitude was increased in increments of 0.5 mA until i) SID occurred, or ii) a troublesome, non-tolerable side effect other than SID occurred, or iii) a maximum amplitude of 10 mA was reached. The occurrence of SID was determined by consensus of two experienced raters (TT (phonetician) and HJ (MD)), based on repetitive naming of the months. In each condition (“stimulation ON”, “stimulation OFF”, monopolar review endpoints), speech and motor symptoms were assessed. Of note, we prematurely stopped the assessment if patients became too exhausted during the assessments to prevent a deterioration of data quality due to poor speech performance (N = 4 contacts).

*Speech assessment* followed a previous study by our group investigating SID in patients with essential tremor and thalamic DBS.^24^ In the present study, patients were asked to enumerate the months’ names and rate their “ability to speak” on a visual analogue scale (VAS) ranging from 0 (normal) to 100 (worst). The examiners were blinded to the patient’s self-ratings. Patients read a well-established German standard text (“Northwind and Sun”) at comfortable reading speed and loudness for intelligibility ratings with additional naïve listeners. Speech data were recorded in a specifically prepared low-noise environment with a headset condenser microphone (AKG 520, mono, 16 bits, 41 kHz) keeping the mouth-to-microphone distance constant. The gain level was not adjusted between the recording sessions and conditions. Before the study procedures, all patients practiced reading the text at least once. To this end, the sentence with the fewest errors and the highest reading fluency was extracted and rated by 15 naïve listeners in an individually randomized order to avoid a potential bias on the ratings. Each stimulus was evaluated on a VAS reaching from 0 (“very poor intelligibility”) to 100 points (“very good intelligibility”). For further analysis, intelligibility ratings per sample were calculated as the mean of all ratings across naïve listeners.

For *motor assessment* the Unified Parkinson’s Disease Rating Scale (UPDRS)-III was employed. Regarding time-efficiency, the complete assessment was only conducted in “stimulation ON” and “stimulation OFF” conditions. During the monopolar review motor assessment took place at an amplitude of 2 mA to ensure the comparability of each contact levels’ efficiency of motor symptom control. If 2 mA were not tolerated, the highest tolerated amplitude was used for motor assessment. Concerning time-efficiency, only items 22 (rigidity), and 23 (finger tapping) were assessed during the monopolar review and summed up as the motor control score. Additionally, presence of ataxia was assessed by asking the patients to point their index finger at the examiner’s index finger back and forth, and the occurrence of any other stimulation-induced side effect was documented (e.g., paresthesia, diplopia, or signs of internal capsule activation) was recorded respectively.

### Localization of DBS Leads and Estimation of Stimulation Volumes

DBS leads were localized with the LEAD-DBS toolbox (www.lead-dbs.org). The detailed preprocessing pipeline has been described previously.^24,25^ In brief, postoperative computed tomographic images (IQon Spectral CT, iCT 256, Brilliance 256, Philips Healthcare, Best, the Netherlands) were linearly coregistered to preoperative magnetic resonance imaging (3T Ingenia, Achieva, 1.5T Ingenia, Philips Healthcare) using advanced normalization tools (ANTs, http://stnava.github.io/ANTs/). Then images were nonlinearly normalized into standard space (ICBM 2009b NLIN asym.) using ANTs and the “effective (low variance)” preset (N = 22), or SPM Shoot or Segment protocols (N = 3) if ANTs resulted in non-optimal fit. DBS leads were automatically reconstructed with the PaCER algorithm,^26^ manually refined, and corrected for postoperative brain shift as implemented in Lead-DBS. The orientation of directional leads was determined with the DiODE algorithm.^27^

For each stimulation setting, precomputed electric fields (FastField) were employed to estimate the spread of the electric field for homogenous tissue with a conductivity of σ = 0.2S/m.^28–30^ The spread of the electric field was modeled in the patient’s native space and then transformed into standard space based on the individual nonlinear transformation. Instead of applying a fixed electric field threshold resulting in a binarized stimulation volume, a novel, more probabilistic stimulation model based on a sigmoidal activation function and published electric field activation thresholds was introduced (see Supplementary Material 1). Of note, the modelled stimulation settings included every investigated 0.5 mA increment during the monopolar review.

### Calculation of Fiber-wise Odds Ratios

We developed a modified fiber-filtering approach for the local mapping of binary outcomes (i.e., SID), based on an approach introduced by Baldermann et al.^31^ Due to the binary nature of our primary outcome (i.e., SID or No SID), we calculated a fiber-wise Odds Ratio for SID for each investigated fiber (see Fig. 1). To do so, we employed a well-established precomputed version of a publicly available PD group connectome (Parkinson’s Progression Markers Initiative (PPMI); N = 90, age 61.4 ±10.42; 28 women; www.ppmi-info.org).^32^ First, the maximal activation probability of each stimulation settings’ sigmoid electric field was determined for each fiber. Only fibers with an arbitrarily chosen summed activation probability ≥ 10 were included for further analysis to control for potential outliers. Second, for each included fiber, an Odds Ratio was determined based on a 2×2 table, including the rater-based distinction between stimulations causing SID and stimulations not causing SID, as well as the likelihood of the fiber being stimulated or not. The likelihood of being stimulated was calculated as the sum of the fiber-wise maximal activation probability of each electric field stimulating the respective fiber. Consecutively the likelihood of not being stimulated was calculated as the sum of 1 minus the fiber-wise maximal activation probability of each electric field stimulating the respective fiber (also see Figure 1B). Haldane-Anscombe correction was applied in case the 2×2 table contained at least one “zero cell”.^33^ These Odds Ratios were not meant to result in significant results but instead informed a model that was tested in a leave-one-out and an out-of-sample validation approach as presented in the following section. All analyses were conducted separately for each hemisphere.

**Figure 1.**
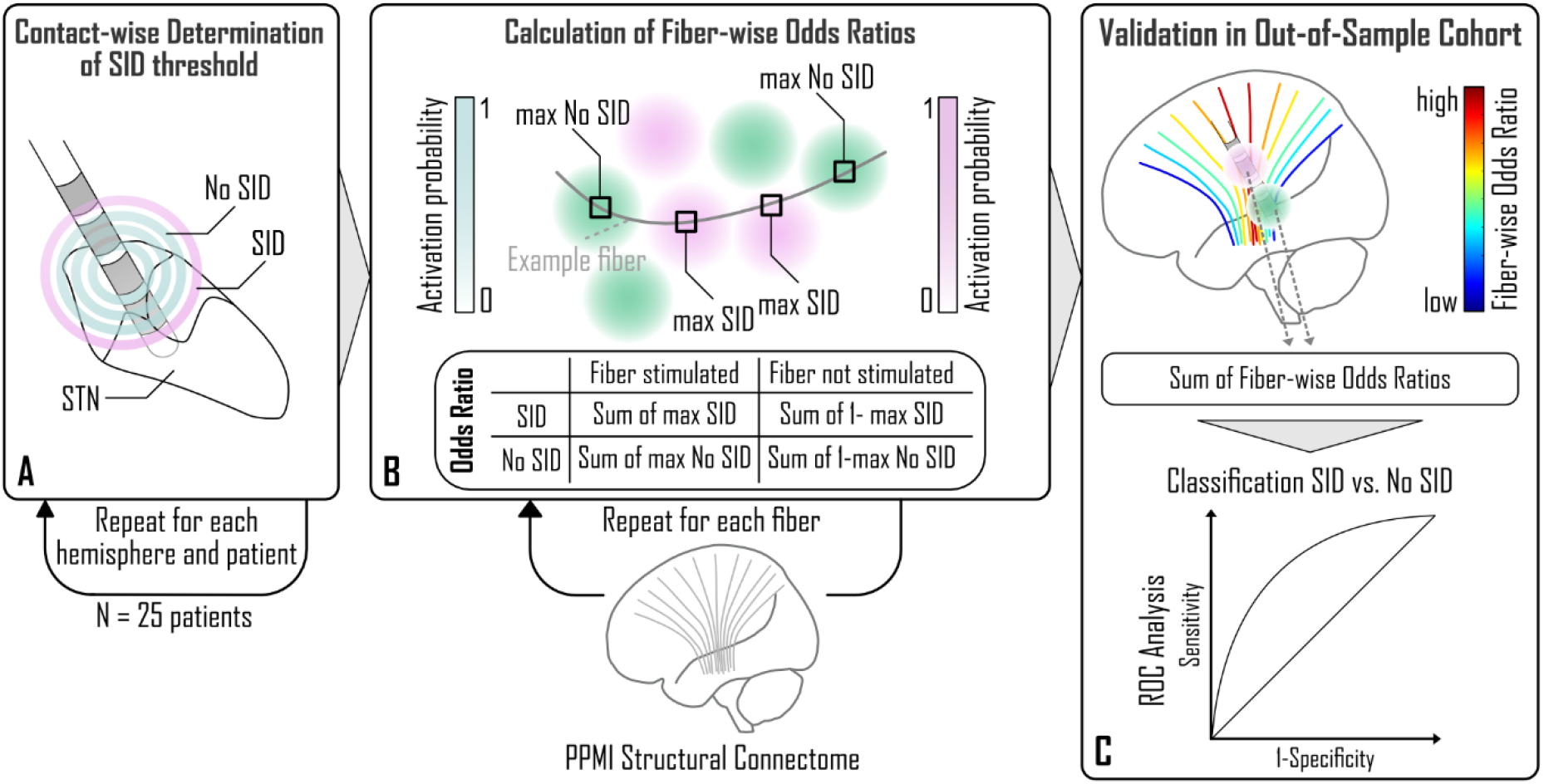
Methods. (A) Thresholds for stimulation-induced dysarthria (SID = pink, No SID = green) were determined in incremental steps of 0.5 mA for each contact in the study cohort. (B) Stimulation spread was modeled as a sigmoid transformation of the electric field distribution, transformed in a common space. Then the Odds Ratio for SID was calculated for each fiber of the employed normative structural connectome based on the fiber-wise maximal activation probability of each stimulation condition. (C) The discriminative performance between “SID” and “No SID” settings of the resulting fiber-wise Odds Ratios was tested in an out-of-sample cohort (N=14), employing a ROC-Analysis. Abbreviations: PPMI = Parkinson’s Progression Markers Initiative, SID = stimulation-induced dysarthria

### ROC-Analysis for Validation of the Data-driven Model

To validate our results, we tested the goodness-of-fit of the estimated fiber-wise Odds Ratio based model to distinguish between stimulation settings causing SID and stimulation settings not-causing SID in i) a leave-one-patient-out approach and ii) an out-of-sample cohort by conducting receiver operating characteristics (ROC) analysis. For *leave-one-patient-out validation*, the model was recalculated 25 times without the data of the respective left-out patient, resulting in the recalculated model serving as the training dataset and the left-out patient data as the test dataset. As the leave-one-patient-out validation in the current study design resembles an artificial setting and as it is impossible to derive a meaningful cutoff value, due to the alternating (leave-one-patient-out) training dataset, an additional out-of-sample validation was conducted. For *out-of-sample validation*, the complete study cohort served as the training dataset. The test dataset comprised 14 randomly chosen independent patients with PD (4 female, age 67.4 ±9.1 years, disease duration 10.7 ±5.0 years) with bilateral STN-DBS who underwent a monopolar review to test for clinical effects and side effects after DBS-surgery as per clinical routine at our center (mean time since surgery 5.2 ±3.8 months). The standardized monopolar review at our center is conducted at least 3 months after surgery and consists of an assessment of motor effects and side-effects of each contact with the contralateral side turned off. The contact order was randomized, and the patient was blinded to the contact selection. Segmented contacts were tested per segment and in omnidirectional (“circular”) mode. Side effect thresholds were assessed under regular medication by increasing the stimulation in steps of 0.5 mA until side-effects occurred or a maximum amplitude of 5 mA was reached. Of note, monopolar reviews were conducted by an independent experienced rater (JNS). Lead reconstruction procedures and stimulation volume calculation were equal to the study cohort, resulting in one sigmoidal transformed electric field for each stimulation setting. For ROC-analysis the predictor variable was calculated as the sum of the weighted Odds Ratios of fibers stimulated by the respective stimulation volume (= fiber-specific maximal activation probability of the respective stimulation volume of the test-dataset*fiber-specific Odds Ratio, generated by the training-dataset). The cutoff value to classify a stimulation volume into “SID” or “No SID” was chosen based on the ROC-curve’s highest Youden’s-index (J_max_). Additionally, the area under the ROC-curve (AUC) as well as the true negative rate (TNR) and the true positive rate (TPR) at J_max_, are reported.

### Statistical Analysis of Secondary Outcomes

If not indicated otherwise, outcomes are reported as mean ± standard deviation. All coordinates are reported as mm-coordinates in MNI space (ICBM 2009b NLIN asym.). To compare additional outcomes, either a t-test or a Wilcoxon rank sum test was applied, depending on the distribution of the data, examined using the Shapiro-Wilk test. If more than two groups were compared, we used the Kruskal-Wallis-Test due to the non-parametric distribution of the data and post-hoc Wilcoxon rank sum tests. If repeated measurements were conducted, the resulting p-values were adjusted by Bonferroni correction. All results are reported to a significance level of p <0.05.

### Data availability

All analyses were conducted with MATLAB 2022a (The MathWorks Inc., Natick, Massachusetts, USA). The resulting fiber-wise Odds Ratio model and the script for the employed sigmoid transformation of the electric field are available via the open science framework (*Link will be added after peer-review*). All other scripts are implemented in LEAD-DBS and were modified by the authors for the needs of the study. Additional clinical and imaging data are available upon reasonable request to the corresponding author.

## Results

### Study Cohort

A total of 25 patients with PD (3 female, age 65.07 ±8.36 years) and bilateral STN-DBS were prospectively recruited, resulting in 50 leads and 200 contact levels investigated. Four contacts were excluded due to abnormal impedances. All participants were right-handed and were included 18 ±12 months after DBS surgery. Lead tip positions did not differ between hemispheres (left hemisphere: x = -11.4 ±1.0 mm, y = -14.7 ±1.1 mm, z = -8.8 ±1.3 mm, right hemisphere: x = 11.5 ±0.9 mm, y = -14.8 ±1.2 mm, z = -8.9 ±1.2 mm; all p >0.05).

### Clinical Outcomes

Clinical outcomes are illustrated in Figure 2. There was no difference in intelligibility ratings (intelligibility “OFF”: 70.07 ±18.37 pts, intelligibility “ON”: 73.79 ±16.72 pts, p = 0.46) or patient-rated VAS “ability to speak” (VAS “OFF” 3.49 ±2.41 pts, VAS “ON” 2.55 ±1.97 pts, p = 0.2). As expected, UPDRS III total scores improved when comparing “stimulation OFF” and “stimulation ON” conditions (UPDRS III total “OFF” 29.12 ±10.4 pts, UPDRS total “ON” 15.75 ±6.32 pts, p <0.001).

**Figure 2.**
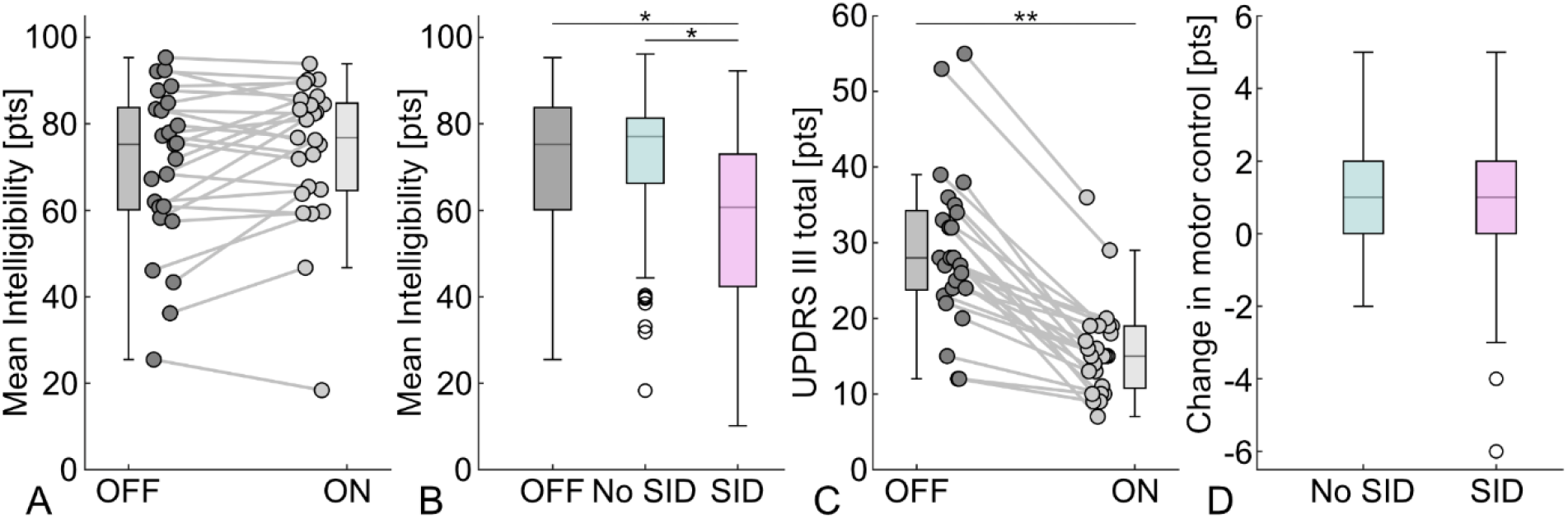
Clinical Outcomes. (A) No difference in intelligibility ratings between “stimulation OFF” and “stimulation ON” conditions. (B) Blinded intelligibility ratings by naïve listeners worsened in stimulation settings classified as causing stimulation-induced dysarthria (SID) compared to stimulation settings not causing SID and “stimulation OFF”. (C) UPDRS III total scores improved in “stimulation ON” settings. (D) Motor symptom control with a stimulation amplitude of 2 mA did not differ between contacts causing SID and contacts not causing SID. *p<0.05, **p<0.001 Abbreviations: UPDRS = Unified Parkinson’s Disease Rating Scale, SID = stimulation-induced dysarthria

As illustrated in Figure 3, SID was evocable at 132 of 196 investigated contacts (N_left_ = 68, N_right_ = 60). The amplitude of the SID threshold did not differ between the hemispheres (SID threshold left: 4.3 ±1.6 mA, SID threshold right: 4.5 ±1.3 mA; p = 0.08). Of note, this SID threshold has to be interpreted per the study design, including an increase of the stimulation amplitude beyond the onset of the first side-effect, if tolerated, to determine the SID threshold. Contact positions for contacts causing SID (x_left_ = -12.4 ±1.3 mm, x_right_ = 12.7 ±1.2 mm; y_left_ = -13.2 ±1.6 mm, y_right_ = -12.8 ±1.8 mm; z_left_ = -6.4 ±2.3 mm, z_right_ = -6.4 ±1.9 mm) and contacts not causing SID (x_left_ = -12.4 ±1.3 mm, x_right_ = 12.6 ±1.3 mm; y_left_ = -13.2 ±1.6 mm, y_right_ = -13.2 ±1.8 mm; z_left_ = -6.4 ±2.3 mm, z_right_ = -6.5 ±2.5 mm) did neither differ on the right nor the left hemisphere (all p >0.05). The position of contacts causing SID did not differ between both hemispheres (all p >0.05). Additionally, there was no correlation between the SID threshold and hemisphere-wise x-, y- or z-coordinates of contacts causing SID (all p >0.05). Accompanying side effects are demonstrated in Figure 3 and were well-balanced between the hemispheres, although muscle contractions tended to accompany SID more often on the right hemisphere (N_left_ = 15, N_right_ = 24). Contacts classified as causing SID did not provide different motor symptom control (1.65 ±1.51 pts) as contacts not causing SID (1.85 ±1.52 pts, p = 0.24). As expected, intelligibility, rated by naïve listeners, was decreased at the SID threshold at contacts causing SID (57.84 ±19.87 pts) in comparison to contacts not causing SID at the highest tolerated amplitude (71.22 ±17.03 pts, p <0.001) or “stimulation OFF” state (70.07 ±18.37 pts, p = 0.012). However, intelligibility did not differ between “stimulation OFF” state and contacts not causing SID at the highest tolerated amplitude (p >0.05). The same results were demonstrated when investigating the patient-rated VAS “ability to speak” (contact levels causing SID: 5.47 ±2.23 pts, “stimulation OFF” 3.49 ±2.41 pts, p_SIDvsOFF_ <0.001, contact levels not causing SID: 4.19 ±2.58 pts, p_SIDvsNOSID_ = 0.004, p_NOSIDvsOFF_ = 0.46).

**Figure 3.**
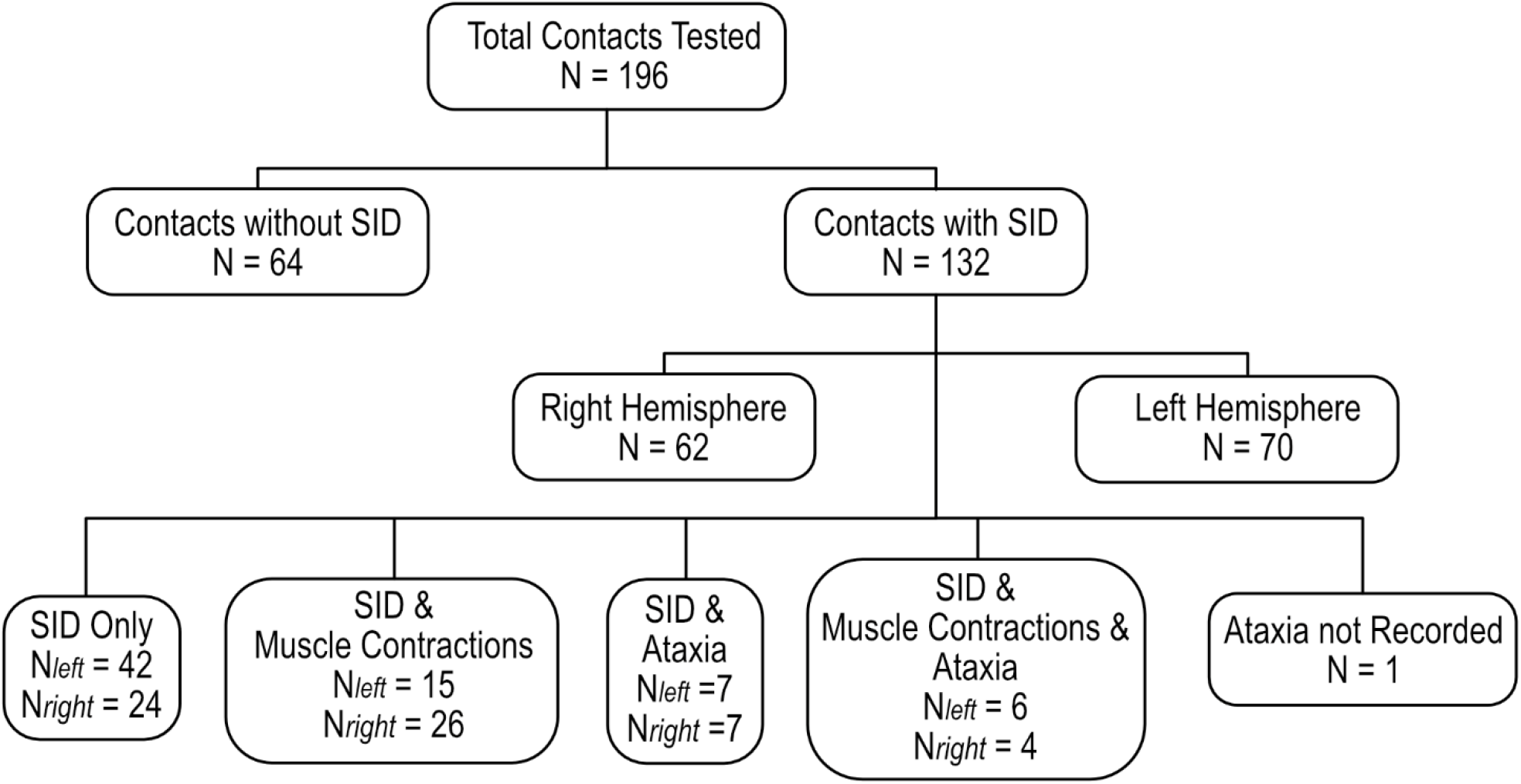
Side effects associated with stimulation-induced dysarthria. A total of 196 contacts were tested, and SID was provoked at 132 contacts. While sometimes appearing as the sole side effect, SID was mainly accompanied by muscle contractions, ataxia, or both. Of note, during one testing session, assessment for ataxia was not performed. Abbreviations: SID = stimulation-induced dysarthria

### Local Mapping of Fiber-wise Odds Ratios for SID

A total of N = 1614 stimulation settings (N_left_ = 798, N_right_ = 816) were included for calculating the fiber-wise Odds Ratios. Fibers with higher Odds Ratios for SID were located both lateral and postero-medial to the STN in both hemispheres (see Figures 4 and 5).

**Figure 4.**
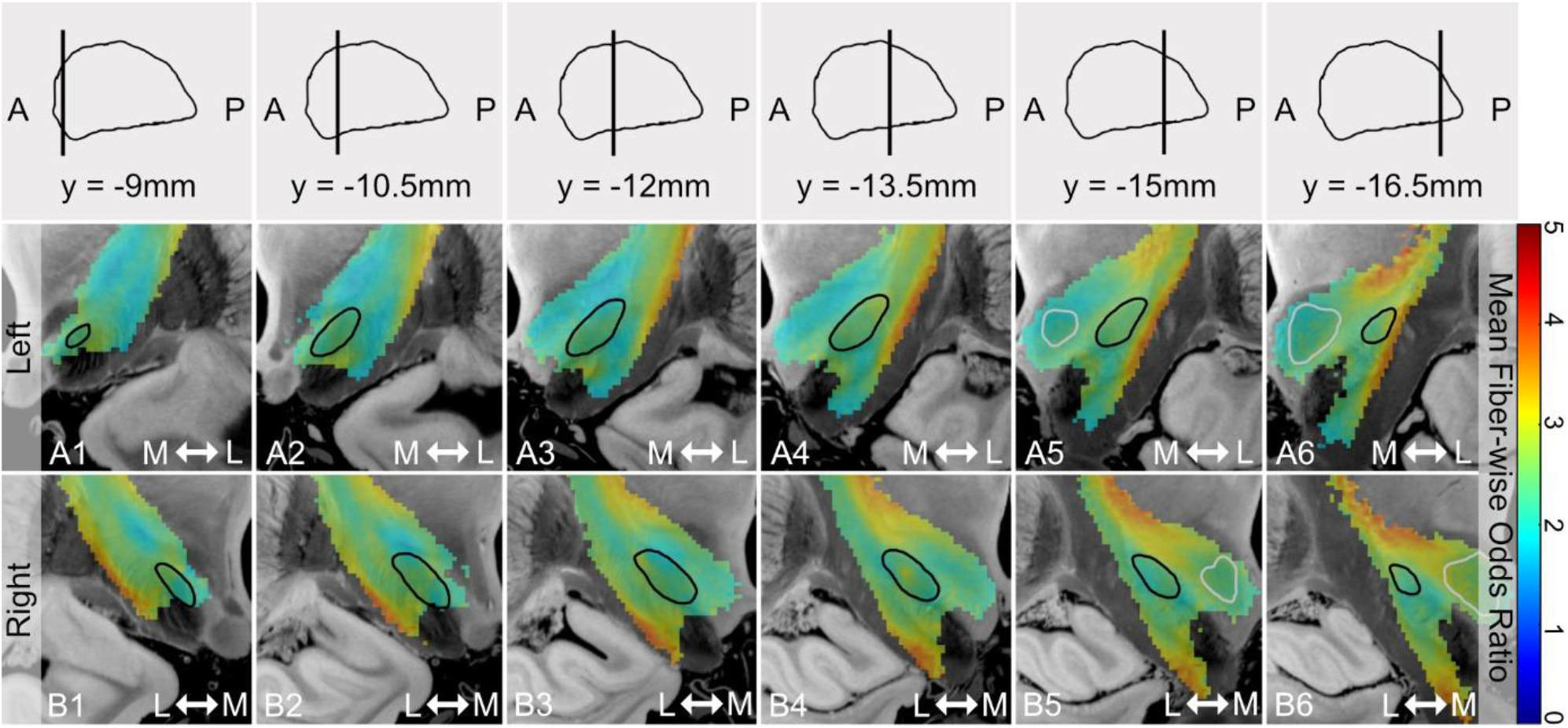
Local Distribution of Fiber-wise Odds Ratios. For illustration, voxels were color-coded by the mean Odds Ratio of all fibers traversing the respective voxel. Voxels with higher mean fiber-wise Odds Ratios are located lateral and postero-medial to the STN (A&B). All maps are superimposed on coronal slices of the BigBrain dataset.^48^ Slice positions in relation to the STN in sagittal view are indicated in the first row. Black outlines indicate the STN, and white outlines the Nucleus ruber.^32^ Abbreviations: A = anterior, L = lateral, M = medial, P = posterior, STN = subthalamic nucleus

**Figure 5.**
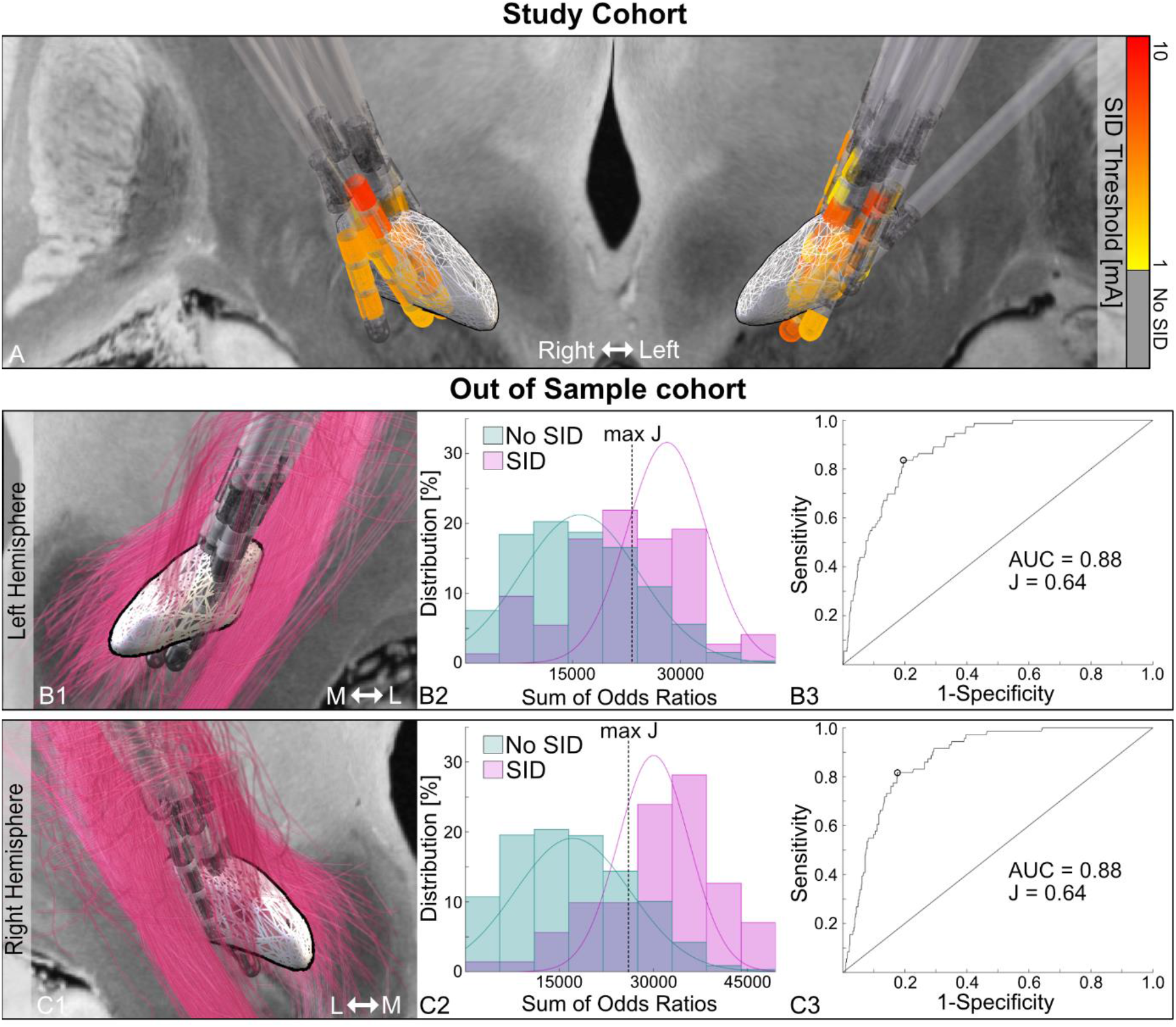
Lead Positions and Out-of-Sample Validation. Lead positions are illustrated in anterior view in relation to the STN in the study cohort (A), and the out-of-sample cohort (B1&C1).^32^ In (A) the contact positions are color-coded by the amplitude to provoke SID, and revealing no clear distributional pattern. In (B1&C1) the leads are shown together with the hemisphere-wise fibers with the 5% highest Odds Ratios. In (B2&C2) histograms illustrate the distribution of the weighted sum of Odds Ratios, calculated based on the study cohort, for the stimulation volumes of the out-of-sample cohort. Stimulation volumes are categorized in “SID” (pink) and “No SID” (green), based on clinical testing. The corresponding ROC-analysis are shown in B3 and C3. Abbreviations: AUC = Area under the curve, J = Youden’s index, L = lateral, M = medial, SID = stimulation-induced-dysarthria, STN = Subthalamic Nucleus; Backdrop: BigBrain Dataset^48^

### Model Validation

ROC-analysis of the leave-one-patient-out validation resulted in an area-under-the-curve (AUC) of 0.75 for the left hemisphere and 0.77 for the right hemisphere. As illustrated in Figure 5, the ROC-analysis with the study cohort as the training dataset and the out-of-sample cohort as the test dataset, resulted in an AUC of 0.88 for the left hemisphere and 0.88 for the right hemisphere. The cutoff weighted sum of fiber-wise Odds Ratios to classify a stimulation volume into “SID” or “No SID” was 23550 for the left hemisphere (J_max_ = 0.64) with a TPR of 83.6%, a TNR of 80.4%, and a precision of 80.9%. For the right hemisphere the cutoff was 26277 (J_max_ = 0.64), resulting in a TPR of 81.7%, a TNR of 82.4%, and a precision of 82.3%.

In a post-hoc analysis we compared this model to the stimulation amplitude as a predictor. ROC-analysis resulted in a similar AUC (AUC_left_ = 0.88, AUC_right_ = 0.86) and a slightly higher TPR at a cutoff of 2.5 mA (J_max-left_ = 0.64, J_max-right_ = 0.60), whereas the TNR was lower (left hemisphere: TPR = 89.0%, TNR = 74.9%, precision = 78.0%; right hemisphere: TPR = 87.3%, TNR = 73.1%, precision = 76.5%). The out-of-sample cohort comprised a total of N = 280 contacts tested, resulting in N = 1960 stimulation settings (N_left_ = 985, N_right_ = 975). SID occurred at N = 144 contacts (N_left_ = 73, N_right_ = 71) at an amplitude of 2.2 ±1.3 mA on the left, and 2.1 ±1.2 mA on the right hemisphere (p_leftvsright_ = 0.57).

## Discussion

This prospective, double-blinded, monocentric study in patients with PD and bilateral STN-DBS demonstrates that stimulation-induced dysarthria (SID) is associated with stimulation spread to fibers lateral and postero-medial to the STN. Furthermore, the data-driven model correctly classified stimulation volumes as causing SID in 83.6% of the cases on the left hemisphere, (AUC = 0.88), and in 81.7% of the cases on the right hemisphere (AUC = 0.88) in an independent out-of-sample cohort. These results are based on the clinical testing of 25 patients with PD and bilateral STN-DBS, resulting in a total of N = 196 contacts tested and N = 1614 stimulation settings included. The finding that contacts causing SID and contacts not causing SID were equally effective regarding motor symptom control underlines the therapy-limiting potential and the need to deepen the pathophysiologic understanding of this clinically relevant side effect (see Fig. 2).

### Structural networks associated with SID

The spatial distribution of fibers with high Odds Ratios in the resulting maps of local fiber-based mapping point toward the involvement of two separate motor networks, the pyramidal tract, as well as cerebello-thalamic fibers, in the pathogenesis of SID, independent of the investigated hemisphere (see Fig. 4 and 5). The essential role of these main motor networks for speech motor control in patients with PD aligns with previous evidence from an FDG-PET study by Pinto et al.^34^ Regarding the origin of stimulation-induced impairment of the speech motor system, the existing literature so far remains heterogeneous. Previous studies reported contacts placed medial and posterior to the STN but not lateral to the STN to be associated with SID, suggesting involvement of cerebello-thalamic fibers.^12–14^ In contrast, other studies found an association between SID and laterally placed contacts, implying an involvement of the pyramidal tract, i.e. stimulation spread to corticobulbar fibers,^15,17^ or both, involvement of the pyramidal tract and cerebello-thalamic fibers.^16,35^ The involvement of these two motor networks in the pathogenesis of SID is also supported by a previous study of our group investigating structural networks associated with SID in thalamic DBS for patients with Essential tremor.^24^ Although there is evidence for the occurrence of distinct phenotypes of speech deterioration after DBS, to date there is only limited evidence for distinct networks, leading to these distinguishable phenotypes.^36^ Additionally, previous studies suggest a crucial role of left-hemispheric stimulation in the pathogenesis of SID.^5,13,37^ Such a hemispheric specialization for SID is not supported by our current data as i) the frequency of SID or the amplitude to provoke SID during clinical testing, ii) the contact position between contacts causing SID and contacts not causing SID, and iii) the resulting local SID maps were not different when comparing both hemispheres.

### Methodological Considerations and Limitations

In the present study, we chose a binary classification into “SID” or “No SID”, based on the consensus of two experienced raters, as the primary outcome parameter. While this dichotomous outcome is not uncommon in the investigation of dysarthria as a stimulation-induced side effect,^15–17^ studies investigating the effects of DBS on the speech motor system often chose intelligibility ratings as primary outcome parameter.^5,7,8,11,14,24^ However, in our experience the change in intelligibility achieved by a particular stimulation condition compared to the “stimulation OFF” condition is prone to be distorted by the occurrence of hypokinetic dysarthria in the “stimulation OFF” condition. The possible stimulation-associated amelioration of hypokinetic dysarthria by DBS can improve the overall intelligibility ratings,^5^ outweighing the worsening of intelligibility caused by the stimulation itself, i.e., SID. Consequently, an improvement in the overall intelligibility under stimulation could be observed, even while SID was detected. Therefore, the binary classification approach (“SID” versus “No SID”) was employed to ensure the capture of SID occurrence as primary outcome of the present study. Of note, overall intelligibility was worse for stimulation settings causing SID than stimulation setting not causing SID and “OFF stimulation” condition (see Fig. 2).

Previous studies investigating the influence of DBS on the speech motor system were limited to comparing “stimulation OFF” to “stimulation ON” conditions or longitudinal assessments of changes in speech after DBS^6–8,11,36,37^ and did not account for stimulation spread,^14,16^ nor did they implement structural connectivity analysis.^13^ We combined a systematic assessment of each contact for the occurrence of SID with a state-of-the-art fiber-based mapping approach, taking the individual stimulation spread of each investigated stimulation setting and its structural connectivity into account.

To estimate the electric field of the stimulation volume, we employed a well-established approach previously used to identify optimal target regions and connectivity profiles in DBS.^24,25,28,38–41^ In the present study, the resulting electric fields were transformed based on a novel introduced sigmoidal activation function, implementing the electric field thresholds for the activation of axons with different diameters as published by Aström et al (see Supp. Material 1).^30^ This approach, in contrast to commonly used binarized stimulation volumes (so called VTAs),^24,39,41^ better reflects both the unknown properties of the underlying white matter and the probabilistic activation characteristics of white matter pathways in which no activation occurs below a certain threshold but the saturation of activation is achieved above a certain threshold, following a sigmoidal activation function.^42^ Another advantage of applying this transformation to the electric field, resulting in electric field values reaching from asymptotic 0 to 1, is that we could derive fiber-wise activation probabilities and fiber-wise non-activation probabilities (= 1-activation probability) for each stimulation volume for our fiber-wise mapping approach. This approach, in the first place, allowed us to calculate weighted fiber-wise Odds Ratios for the binary outcome SID. Although it has recently been shown that using weighted stimulation volumes might explain slightly more variance in clinical outcomes than using classic binary stimulation volumes, modelling an all-or-nothing activation, the theoretical concept employed in the present study remains a simplification and neglects factors like, e.g., fiber orientation.^25,43^ To combine the weighted stimulation volumes of all participants into one joint data-driven model, stimulation volumes were normalized to a commonly used standard space (ICBM 2009b NLIN asym.). Although this approach neglects some inter-individual heterogeneity the employed state-of-the-art normalization algorithms implemented in LEAD-DBS, performed similarly to manual expert segmentations.^44^

In contrast to previous local mapping studies, mainly calculating data-driven models on a voxel level, we employed a structural normative disease-matched group connectome to derive a fiber-based local mapping approach. The rationale behind this approach was the hypothesized white-matter origin of the primary outcome (SID) and the expected enhanced spatial information by implementing the structural connectivity between the included stimulation volumes. This allows the model to be informed simultaneously by several non-spatially overlapping stimulation volumes. This is of particular importance when considering the relatively low rate of SID occurrence in comparison to continuously measurable outcomes, e.g., motor symptom control, widely used as an outcome measure in previous voxel-based mapping studies.^39^ For comparison, when repeating the analysis on a purely voxel-based level, the local distribution pattern of SID occurrence was still visible, but less contrasted, indicating a worse signal-to-noise ratio (see Supplementary Fig. 2). Importantly, this does not indicate a general superiority of a fiber-wise versus voxel-wise DBS mapping approaches, and of course the interpretation of these results underlies the same limitations.^45^ However, it supports the use of fiber-wise mapping in cases where a white-matter origin of the respective outcome is likely. Of note, the employed normative structural connectome, based on data acquired in patients with PD in the PPMI project, does not account for individual structural connectivity and might be prone to false-positive connections.^46^ On the one hand, normative connectomes, as used in this study, comprise a large dataset, leading to high signal-to-noise levels and state-of-the-art data quality. Several recent studies, investigating DBS effects have validated the use of this normative connectome in PD.^40,47^ Conversely, applying different normative connectomes might lead to distorted results based on the data and methods used to generate them.

Further, we validated the resulting data-driven model and demonstrated high accuracy in an independent out-of-sample cohort comprising 14 patients with PD and bilateral STN-DBS and N = 1960 stimulation settings in omnidirectional and directional conditions (see Supp. Material 2). While the amplitude as the sole predictor of SID (and other stimulation-induced side effects) performed similarly to the data-driven model in a ROC-analysis at a cutoff value of 2.5 mA to classify stimulation settings as causing SID, it does not provide insight into the local current distribution and thus pathoanatomical correlates of side-effects. The finding, that the ROC-analysis suggested a slightly better fit of the data-driven model in the out-of-sample cohort than in the leave-one-patient-out cohort is most likely attributed to the artificial test setting in the original cohort with an increase of the amplitude beyond the onset of stimulation-induced side effects to elicit SID whenever tolerated. In contrast, clinical testing in the out-of-sample cohort only implemented the determination of side effect thresholds, defined as the onset of any side effect – so SID was only encountered on contacts closer to the underlying fibers.

## Conclusion

The present study provides evidence that both cerebello-thalamic fibers and the pyramidal tract are involved in the pathogenesis of stimulation-induced dysarthria in patients with PD and STN-DBS, independent of the investigated hemisphere. Future studies should focus on the identification of possible subtypes of SID and their respective association with the stimulation of distinct motor networks, the combination of network analysis of SID across target points, and the validation and implementation of these data-driven models in imaging-guided programming strategies to improve the clinical care of patients with DBS.

## Data Availability

All data produced in the present study are available upon reasonable request to the authors

## Abbreviations

ANTs: advanced normalization tools
AUC: Area-under-the curve
CAPSIT-PD: Core assessment program for surgical interventional therapies in Parkinson’s disease
DBS: Deep Brain Stimulation
FDG-PET: fluorodeoxyglucose (FDG)-positron emission tomography (PET)
J: Youden’s-index
PD: Parkinson’s Disease
ROC: Receiver operating characteristics
PPMI: Parkinson’s Progression Markers Initiative
SD: standard deviation
SID: stimulation-induced dysarthria
STN: Subthalamic Nucleus
TPR: True positive rate
TNR: True negative rate
UK: United Kingdom
UPDRS: Unified Parkinson’s Disease Rating Scale
VAS: Visual Analogue Scale

## Acknowledgements

We thank our patients for study participation and the naïve listeners for their time and commitment to supporting our study.

JPS and TAD were funded by the Cologne Clinician Scientist Program (CCSP)/ Faculty of Medicine/ University of Cologne. Funded by the German Research Foundation (DFG, FI 773/15-1). JH was supported by the Koeln Fortune Program/ Faculty of Medicine, University of Cologne. JCB and GRF were funded by the Deutsche Forschungsgemeinschaft (DFG, German Research Foundation): Project-ID 431549029 – SFB 1451. Additionally, JCB was funded by the Else Kröner-Fresenius-Stiftung (grant number 2022_EKES.23) and receives funding from the German Research Foundation (CRC-1451). DM and TT were supported by the Deutsche Forschungsgemeinschaft (DFG, German Research Foundation): Project-ID 281511265 - SFB1252.

Data used in the preparation of this article were obtained from the Parkinson’s Progression Markers Initiative (PPMI) database (www.ppmi-info.org/data). For up-to-date information on the study, visit www.ppmi-info.org. PPMI—a public-private partnership—is funded by the Michael J. Fox Foundation for Parkinson’s Research and funding partners, see http://www.ppmi-info.org/fundingpartners.

## Author Contributions

1) conception and design of the study: HJ, JPS, TT, TAD, MTB 2) acquisition and analysis of data: HJ, JPS, JH, TT, JNS, TAD or 3) drafting a significant portion of the manuscript or figures: HJ, JPS, IRF, JQ, JCB, DM, GRF, VVV, TAD, MTB

## Supplementary

### Supplementary Material 1 – Activation model

Most previous publications using probabilistic stimulation mapping are based on binarized stimulation volumes (so called volume of tissue activated, VTA). Those are commonly derived by estimating the electric field and then applying a cutoff value of, e.g., 0.2 V/mm. Voxels with electric field values above this cutoff have a 100 % activation probability, while voxels with electric field values below this cutoff have a 0% activation probability. However, cutoff values are not well defined and often chosen, at least to some degree, arbitrarily.

In 2014, Aström et al. published electric field thresholds for the activation of simulated axons with different diameters and for various stimulation settings.^30^ Since the properties of the axons responsible for stimulation-induced dysarthria are unknown, we decided to exchange the classic binarized VTA approach with a more probabilistic approach for this study.^30^

Based on the electric field thresholds published by Aström et al., we derived a sigmoidal activation function (see Supplementary Figure 1). The function ranged from 0 % activation probability to 100 %. The function was fitted in a way that a 5 % activation probability was reached at an electric field of 0.061 V/mm (lowest published estimate for axons with a diameter of 7.5 μm), while a 95 % activation probability was reached at an electric field of 0.351 V/mm (highest published estimate for axons with a diameter of 2.5 μm). The resulting function meant that a 50 % activation probability was reached at an electric field of 0.206 V/mm - close to the commonly used threshold of 0.2 V/mm in many publications using binarized VTAs.

**Supplementary Figure 1.**
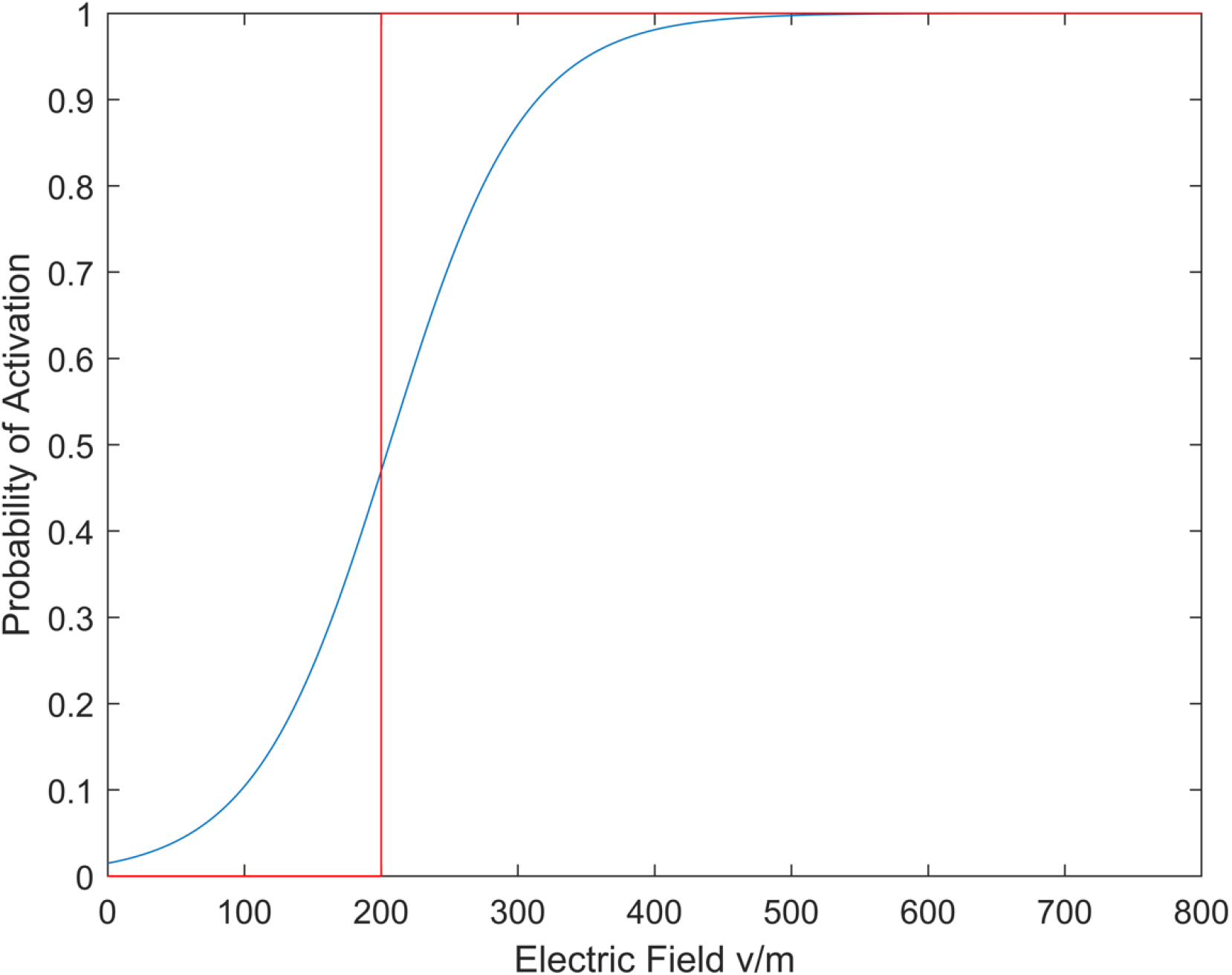
Sigmoid Activation Function. Illustration of the employed sigmoid activation function (blue) and a standard binary activation function (red).

**Supplementary Figure 2.**
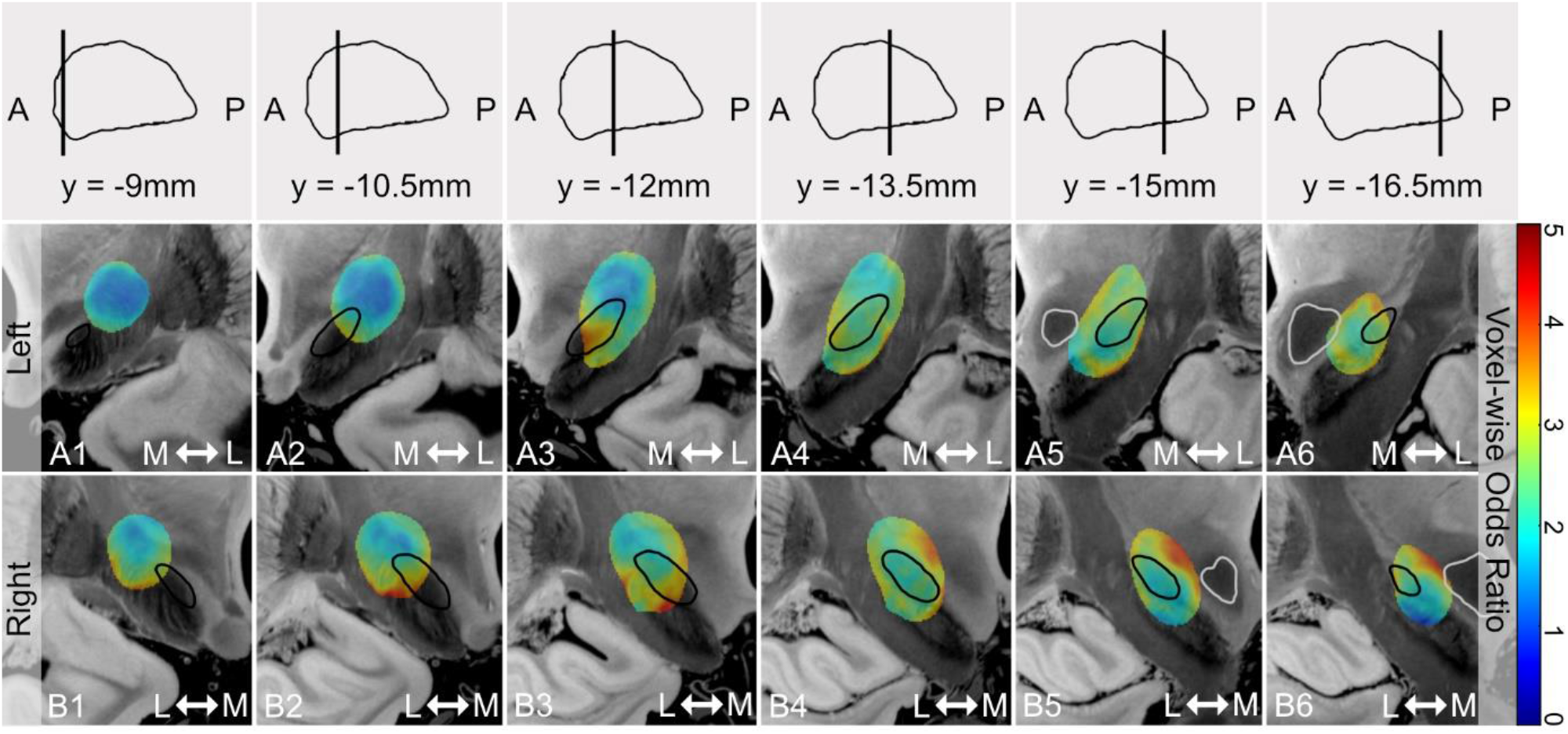
Local Distribution of Voxel-wise Odds Ratios. Voxel-wise Odds Ratios were calculated based on the same approach introduced in the main analysis, with the difference that the Odds Ratios were calculated as per voxel instead of per fiber. The resulting Odds Ratio map is superimposed on coronal slices of the BigBrain dataset with slice positions in relation to the STN in the sagittal view indicated in the first row.^48^ Although a similar pattern of the local distribution of higher Odds Ratios as compared to the fiber-based approach appears, the fiber-based approach provides a better spatial resolution, also accounting for non-spatial overlap of structurally connected stimulation volumes, resulting in a higher signal-to-noise ratio and an overall better interpretability of the results. Black outlines indicate the STN, and white outlines the Nucleus ruber.^32^ Abbreviations: A = anterior, L = lateral, M = medial, P = posterior, STN = Subthalamic nucleus

## References

1. Deuschl G, Schade-Brittinger C, Krack P, et al. A randomized trial of deep-brain stimulation for Parkinson’s disease. N Engl J Med 2006;355(9):896–908.

2. Schuepbach WMM, Rau J, Knudsen K, et al. Neurostimulation for Parkinson’s Disease with Early Motor Complications. New England Journal of Medicine 2013;368(7):610–622.

3. Darley FL, Aronson AE, Brown JR. Motor speech disorders. WB Saunders Company; 1975.

4. Ho AK, Iansek R, Marigliani C, et al. Speech impairment in a large sample of patients with Parkinson’s disease. Behav Neurol 1999;11(3):131–137.

5. Tripoliti E, Zrinzo L, Martinez-Torres I, et al. Effects of subthalamic stimulation on speech of consecutive patients with Parkinson disease. Neurology 2011;76(1):80– 86.

6. Tsuboi T, Watanabe H, Tanaka Y, et al. Distinct phenotypes of speech and voice disorders in Parkinson’s disease after subthalamic nucleus deep brain stimulation. J Neurol Neurosurg Psychiatry 2015;86(8):856–864.

7. Tsuboi T, Watanabe H, Tanaka Y, et al. Early detection of speech and voice disorders in Parkinson’s disease patients treated with subthalamic nucleus deep brain stimulation: a 1-year follow-up study. J Neural Transm (Vienna) 2017;124(12):1547– 1556.

8. Tanaka Y, Tsuboi T, Watanabe H, et al. Longitudinal Speech Change After Subthalamic Nucleus Deep Brain Stimulation in Parkinson’s Disease Patients: A 2-Year Prospective Study. J Parkinsons Dis 2020;10(1):131–140.

9. Aldridge D, Theodoros D, Angwin A, Vogel AP. Speech outcomes in Parkinson’s disease after subthalamic nucleus deep brain stimulation: A systematic review. Parkinsonism Relat Disord 2016;33:3–11.

10. Brabenec L, Mekyska J, Galaz Z, Rektorova I. Speech disorders in Parkinson’s disease: early diagnostics and effects of medication and brain stimulation. J Neural Transm (Vienna) 2017;124(3):303–334.

11. Pinto S, Nebel A, Rau J, et al. Results of a Randomized Clinical Trial of Speech after Early Neurostimulation in Parkinson’s Disease. Mov Disord 2022;

12. Tripoliti E, Zrinzo L, Martinez-Torres I, et al. Effects of contact location and voltage amplitude on speech and movement in bilateral subthalamic nucleus deep brain stimulation. Mov Disord 2008;23(16):2377–2383.

13. Aström M, Tripoliti E, Hariz MI, et al. Patient-specific model-based investigation of speech intelligibility and movement during deep brain stimulation. Stereotact Funct Neurosurg 2010;88(4):224–233.

14. Fenoy AJ, McHenry MA, Schiess MC. Speech changes induced by deep brain stimulation of the subthalamic nucleus in Parkinson disease: involvement of the dentatorubrothalamic tract. J Neurosurg 2017;126(6):2017–2027.

15. Mahlknecht P, Akram H, Georgiev D, et al. Pyramidal tract activation due to subthalamic deep brain stimulation in Parkinson’s disease. Mov Disord 2017;32(8):1174–1182.

16. Strotzer QD, Kohl Z, Anthofer JM, et al. Structural Connectivity Patterns of Side Effects Induced by Subthalamic Deep Brain Stimulation for Parkinson’s Disease. Brain Connect 2022;12(4):374–384.

17. Prent N, Potters WV, Boon LI, et al. Distance to white matter tracts is associated with deep brain stimulation motor outcome in Parkinson’s disease. J Neurosurg 2019;1–10.

18. Dembek TA, Reker P, Visser-Vandewalle V, et al. Directional DBS increases side-effect thresholds-A prospective, double-blind trial. Mov Disord 2017;32(10):1380– 1388.

19. Steffen JK, Reker P, Mennicken FK, et al. Bipolar Directional Deep Brain Stimulation in Essential and Parkinsonian Tremor. Neuromodulation 2020;23(4):543– 549.

20. Barbe MT, Dembek TA, Becker J, et al. Individualized current-shaping reduces DBS-induced dysarthria in patients with essential tremor. Neurology 2014;82(7):614– 619.

21. Fabbri M, Natale F, Artusi CA, et al. Deep brain stimulation fine-tuning in Parkinson’s disease: Short pulse width effect on speech. Parkinsonism & Related Disorders 2021;87:130–134.

22. Fabbri M, Zibetti M, Ferrero G, et al. Is lowering stimulation frequency a feasible option for subthalamic deep brain stimulation in Parkinson’s disease patients with dysarthria? Parkinsonism Relat Disord 2019;64:242–248.

23. Defer G-L, Widner H, Marié R-M, et al. Core assessment program for surgical interventional therapies in Parkinson’s disease (CAPSIT-PD). Movement Disorders 1999;14(4):572–584.

24. Petry-Schmelzer JN, Jergas H, Thies T, et al. Network Fingerprint of Stimulation-Induced Speech Impairment in Essential Tremor. Annals of Neurology 2021;89(2):315–326.

25. Horn A, Li N, Dembek TA, et al. Lead-DBS v2: Towards a comprehensive pipeline for deep brain stimulation imaging. Neuroimage 2019;184:293–316.

26. Husch A V Petersen M, Gemmar P, et al. PaCER - A fully automated method for electrode trajectory and contact reconstruction in deep brain stimulation. Neuroimage Clin 2018;17:80–89.

27. Dembek TA, Hellerbach A, Jergas H, et al. DiODe v2: Unambiguous and Fully-Automated Detection of Directional DBS Lead Orientation. Brain Sciences 2021;11(11):1450.

28. Horn A, Reich M, Vorwerk J, et al. Connectivity Predicts deep brain stimulation outcome in Parkinson disease. Ann Neurol 2017;82(1):67–78.

29. Baniasadi M, Proverbio D, Gonçalves J, et al. FastField: An open-source toolbox for efficient approximation of deep brain stimulation electric fields. NeuroImage 2020;223:117330.

30. Astrom M, Diczfalusy E, Martens H, Wardell K. Relationship between neural activation and electric field distribution during deep brain stimulation. IEEE Trans Biomed Eng 2015;62(2):664–672.

31. Baldermann JC, Melzer C, Zapf A, et al. Connectivity Profile Predictive of Effective Deep Brain Stimulation in Obsessive-Compulsive Disorder. Biol Psychiatry 2019;85(9):735–743.

32. Ewert S, Plettig P, Li N, et al. Toward defining deep brain stimulation targets in MNI space: A subcortical atlas based on multimodal MRI, histology and structural connectivity. Neuroimage 2018;170:271–282.

33. Weber F, Knapp G, Ickstadt K, et al. Zero-cell corrections in random-effects meta-analyses. Res Synth Methods 2020;11(6):913–919.

34. Pinto S, Thobois S, Costes N, et al. Subthalamic nucleus stimulation and dysarthria in Parkinson’s disease: a PET study. Brain 2004;127(Pt 3):602–615.

35. Strotzer QD, Anthofer JM, Faltermeier R, et al. Deep brain stimulation: Connectivity profile for bradykinesia alleviation. Ann Neurol 2019;85(6):852–864.

36. Lange F, Eldebakey H, Hilgenberg A, et al. Distinct phenotypes of stimulation-induced dysarthria represent different cortical networks in STN-DBS. Parkinsonism Relat Disord 2023;109:105347.

37. Tripoliti E, Limousin P, Foltynie T, et al. Predictive factors of speech intelligibility following subthalamic nucleus stimulation in consecutive patients with Parkinson’s disease. Mov Disord 2014;29(4):532–538.

38. Li N, Baldermann JC, Kibleur A, et al. A unified connectomic target for deep brain stimulation in obsessive-compulsive disorder. Nat Commun 2020;11(1):3364.

39. Dembek TA, Barbe MT, Åström M, et al. Probabilistic mapping of deep brain stimulation effects in essential tremor. NeuroImage: Clinical 2017;13:164–173.

40. Irmen F, Horn A, Mosley P, et al. Left Prefrontal Connectivity Links Subthalamic Stimulation with Depressive Symptoms. Ann Neurol 2020;87(6):962–975.

41. Petry-Schmelzer JN, Krause M, Dembek TA, et al. Non-motor outcomes depend on location of neurostimulation in Parkinson’s disease. Brain 2019;142(11):3592– 3604.

42. Neudorfer C, Butenko K, Oxenford S, et al. Lead-DBS v3.0: Mapping deep brain stimulation effects to local anatomy and global networks. NeuroImage 2023;268:119862.

43. Anderson DN, Duffley G, Vorwerk J, et al. Anodic stimulation misunderstood: preferential activation of fiber orientations with anodic waveforms in deep brain stimulation. J Neural Eng 2019;16(1):016026.

44. Ewert S, Horn A, Finkel F, et al. Optimization and comparative evaluation of nonlinear deformation algorithms for atlas-based segmentation of DBS target nuclei. Neuroimage 2019;184:586–598.

45. Dembek TA, Barbe MT, Åström M, et al. Probabilistic mapping of deep brain stimulation effects in essential tremor. Neuroimage Clin 2016;13:164–173.

46. Petersen MV, Mlakar J, Haber SN, et al. Holographic Reconstruction of Axonal Pathways in the Human Brain. Neuron 2019;104(6):1056-1064.e3.

47. Lofredi R, Auernig GC, Irmen F, et al. Subthalamic stimulation impairs stopping of ongoing movements. Brain 2021;144(1):44–52.

48. Edlow BL, Mareyam A, Horn A, et al. 7 Tesla MRI of the ex vivo human brain at 100 micron resolution. Sci Data 2019;6(1):244.

